# Efficacy of oral Labetalol vs Nifedipine in the management of severe hypertension in pregnancy. A Randomized Controlled Trial

**DOI:** 10.1101/2025.05.06.25327088

**Authors:** Obinna C Eze, Akintunde O Fehintola, Akinfaderin D. Adeyinka, Clement A Adepiti, Ibraheem O Awowole, Adebimpe O Ijarotimi, Babalola E. Olajide, Ajiboye D Akinyosoye, Abdur-Rahim F Zainab, Ayegbusi E Oluwole, Ernest O Orji

## Abstract

Hypertensive disorders in pregnancy (HDP) are leading causes of pregnancy-related morbidity and mortality. International guidelines have recommended the use of intravenous hydralazine or Labetalol in the control of severe HDP. Oral Nifedipine or Labetalol have not been permitted as first-line in managing severe HDP even in low-resource settings where skill for intravenous access is not readily available. This study aimed to determine and compare the efficacy of oral Labetalol versus Nifedipine retard in achieving adequate blood pressure (BP) control among pregnant women with severe Hypertension at Obafemi Awolowo University Teaching Hospitals Complex, Ile-Ife.

This was a single-center, open-labeled, randomized controlled trial where 176 eligible pregnant women with severe BP, 30 minutes after Magnesium Sulfate administration, were randomized into Nifedipine and labetalol groups. Each group received either oral Labetalol or Nifedipine retard; 200mg and 20mg, respectively. The second and third doses were subsequently administered after an hour of the initial dose if adequate BP control was not achieved. The mean BP control, doses used, time of achieving blood pressure control, adverse outcomes, and the Effect of co-administration of both oral antihypertensives one hour after the third dose were assessed and compared accordingly. Data was analyzed using SPSS version 26. The efficacy of oral Labetalol versus Nifedipine in controlling BP among the participants was tested using a student’s t-test and chi-square. P-value < 0.05 was taken to be statistically significant.

There was a statistically significant BP control with co-administration of both anti-hypertensive after single agent doses, 70 (86.4%) vs. 65 (80.2%) for the labetalol and Nifedipine groups, respectively (p= 0.003). Participants in the labetalol group used fewer doses cumulatively, with statistical differences at P values of 0.015 and 0.0001 at the first and second doses, respectively. Participants in the labetalol group achieved BP control within a shorter period with minimal adverse outcomes.

Both oral antihypertensives were effective in controlling severe HDP. Oral Labetalol controlled severe Hypertension better with fewer doses, shorter duration, and fewer adverse outcomes; thus, it should be preferred.

## Introduction

Hypertension is the most common medical disorder of pregnancy, complicating one in ten pregnancies [1, 2]. Hypertension in pregnancy may be chronic (predating pregnancy or diagnosed before 20 weeks of pregnancy) or de novo (either pre-eclampsia or pregnancy-induced Hypertension [1, 2]. Chronic Hypertension is associated with adverse maternal and fetal outcomes. It is best managed by tightly controlling maternal blood pressure to systolic blood pressure (SBP) - 110–140 mmHg and diastolic blood pressure (DBP) 80-90 mmHg, monitoring fetal growth, and repeatedly assessing for the development of pre-eclampsia and maternal complications. Pre-eclampsia is a multisystemic disorder characterized by a new onset of Hypertension with systolic blood pressure (SBP) ≥ 140mmHg and/or diastolic blood pressure (DBP) ≥ 90mmHg accompanied by significant proteinuria and/or maternal end-organ dysfunction, and/or uteroplacental insufficiency after 20 weeks of gestation in a previously normotensive woman which resolves within 6 weeks after delivery [1, 2]. Pregnancy-induced Hypertension is the onset of Hypertension after 20 weeks in the absence of proteinuria and completely resolves before the 6^th^ week of puerperium. Pregnancy-induced Hypertension and pre-eclampsia contribute significantly to maternal mortality, premature birth, intra-uterine growth restriction, and perinatal mortality.

Labetalol is a nonselective β-blocker with vascular α_1_-receptor blocking capabilities. Oral Labetalol has a reduced relative bioavailability of 25% compared to intravenous (IV) labetalol. It also has delayed onset of action (20 minutes as against 2-5 minutes for intravenous Labetalol) and peak effect of 1-4 hours as against 15 minutes for IV labetalol [3–5]. Intravenous Labetalol needs exceptional preservation at temperatures between 4°C and 25°C for maximum efficacy. This may not be feasible in rural settings because of poor and inadequate power supply, lack of refrigerators, and lack of skill to insert intravenous lines. Thus, there is a need to encourage and promote the use of oral antihypertensives in such settings.

Nifedipine is a calcium channel blocker that causes peripheral arterial vasodilation to reduce blood pressure. It is 92-98% protein bound. Its onset of action is within 20-30 minutes and has a half-life of about 4-7 hours. Long-acting preparations are preferred, although obstetric experience with short-acting has been favorable [6]. Obstetric experience prefers the use of short-acting tablets because of their rapid onset of action (buccal-10-15 minutes; oral, 30 – 45 minutes) and peak effect (buccal, 30 minutes, oral, 60 minutes) with a half-life of about 4-7 hours. These preparations cause rapid vasodilatation followed by reflex sympathetic activation, resulting in side effects such as headaches, palpitations, and flushing, which may have adverse maternal and fetal outcomes. Nifedipine Retard is readily absorbed when taken orally with an onset of action of 1-2 hours after administration which is delayed compared to the immediate release of Nifedipine, which is 30 – 60 minutes. The peak concentration time is 6-12 hours, the half-life is 7-12 hours, and the duration of action is about 24 hours. Extended-release preparations have up to 89% bioavailability relative to the immediate-release formulation [7, 8].

Common adverse effects of Nifedipine include flushing, peripheral edema, dizziness, and headache. Tolerance is better with extended-release preparations than with immediate-release preparations. Nifedipine retard was chosen for this study because of its relatively favourable pharmacodynamics and pharmacokinetics [9].

Orally administered Nifedipine and Verapamil do not seem to pose teratogenic risks to fetuses exposed in the first trimester [10]. Most investigators have focused on the use of Nifedipine, although there are reports that nicardipine [11–13], Isradipine [14], and Verapamil [11] can be used with some safety concerns.

One study has shown the efficacy and safety of long-acting oral Nifedipine in pregnant patients with severe Hypertension [15], and given possible untoward fatal effects of short-acting sublingual Nifedipine advocated the use of the long-acting preparation [16, 17].

A concern with the use of calcium channel antagonists for BP control in Pre-eclampsia has been the concomitant use of magnesium sulfate to prevent seizures. Drug interactions between Nifedipine and magnesium sulfate were reported to cause neuromuscular blockade [18], myocardial depression, or circulatory collapse in some cases [19, 20]. In practice and a recent evaluation [21], these medications are commonly used together without increased risk [22, 23]. It is heat stable, comes only in tablet form, and has been recommended by international guidelines for treating severe Hypertension in pregnancy [15, 24].

There is consensus that severe Hypertension in pregnancy, defined as ≥160/110 mm Hg, requires treatment because these women are at an increased risk of intracerebral hemorrhage and that treatment decreases the risk of maternal death [1, 25]. Intravenous hydralazine and Labetalol have been recommended by the International Society for the Study of Hypertension in Pregnancy (ISSHP) guidelines for the management of severe Hypertension in pregnancy [26]. Those with hypertensive encephalopathy, hemorrhage, or eclampsia require treatment with these parenteral agents to lower mean arterial pressure (2/3 diastolic +1/3 systolic BP) by 25% over minutes to hours and then to further lower BP from 160/100 mm Hg over subsequent hours [1]. In treating severe Hypertension, it is essential to avoid hypotension because the degree to which placental blood flow is autoregulated is not established, and aggressive lowering may cause fetal distress. Oral anti-hypeertensive has been found to have less risk of aggressive lowering of BP and has the advantage of prompt initiation of therapy in these women.

In women with pre-eclampsia, consideration should be given to initiating parenteral agents for treatment of acute severe Hypertension at lower doses or with oral antihypertensives because their intravascular volume may be depleted and at increased risk of hypotension [26, 27]. Oral Labetalol or Nifedipine can be safely used in low-resource settings, as the International Society for the Study of Hypertension in Pregnancy recommends because they are readily available and easy to administer [26]. Furthermore, there is a paucity of trained staff with skills needed to site intravenous lines for administering intravenous medications, with fewer centers having gadgets for monitoring fetal and maternal well-being. Despite the limitations regarding the use of parenteral anti-hypertensive in our settings, there is not enough evidence to recommend oral antihypertensives as the first-line drugs in the management of severe Hypertension in pregnancy, hence the drive for this study to assess and compare the efficacy of both oral Labetalol and Nifedipine.

## Methods

### Study location and period

The study was conducted in the Department of Obstetrics, Gynaecology, and Perinatology at the Obafemi Awolowo University Teaching Hospitals Complex (OAUTHC), Ile-Ife, between May 2023 and March 2024. The hospital has two major arms: The Ife Hospital Unit (IHU) Ile-Ife and Wesley Guild Hospital (WGH) Ilesha, both in Osun state. These hospitals serve as referral centers to primary and secondary healthcare providers in Osun State, Ekiti State, Ondo State, and some parts of Oyo State. The two arms of the hospital were used for the study. Both arms of the hospital have a combined annual antenatal clinic booking rate of 2000 per year, with an incidence of severe Hypertension in pregnancy of 3.8%.[28]

### Study design

The study was a single-center, open-label, randomized controlled trial.

### Study Population

The study population comprised all consenting pregnant women with severe Hypertension aged 15-49 years at a gestational age of > 28 weeks, presenting at the antenatal clinic, labor ward outpatient section, and emergency units.

### Inclusion Criteria

All eligible consenting pregnant women aged 15-49 years at a gestational of 28 weeks and above with severe Hypertension in pregnancy.

### Exclusion Criteria

The study excluded pregnant women who were actively wheezing or asthmatic, pregnant women with comorbidities, e.g., known heart disease patients, diabetes mellitus, chronic renal disease, or pregnant women with eclampsia, twin pregnancy, clinical symptoms and signs suggestive of heart disease, and those who react to both antihypertensives. Pregnant women who refused to participate in the study were also excluded.

### Sample Size Determination

This recruited 176 participants for this study using the formula for superiority randomized controlled trial to provide the sample size for each of the two groups [29, 30]

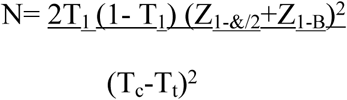

Where N = calculated sample size per group, the probability of success with each drug use is T_C_ and T_t._ The average success rate T_1_ = (T_c_ x T_t_)/2= (0.625+0.75)/2=0.6875 for each person in the control and test groups. The sample size was estimated based on prior knowledge of the primary outcome of published data by Easterling et al., which assumed a success rate of 75% (0.75) with Nifedipine T_t_ and 62.5% (0.625) with labetalol T_C_ [3] and confidence interval limit at 95% with p-value <0.05, power= 1-B (Type 11 error) at 80%-1-0.8=0.2. Z value at 0.2=0.842

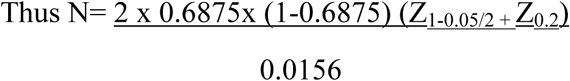

Where Z_1-0.05/2_=Z_0.975_=0.8531, (T_c_-T_t_)^2^=0.0156.

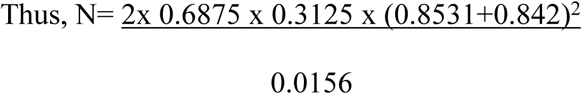

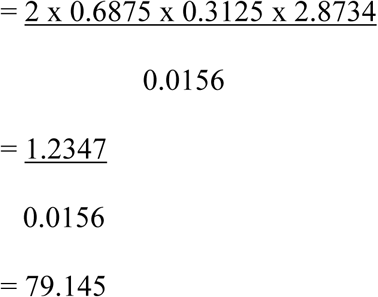

80 in each group, making a total of 160 participants Considering an attrition rate of 10% of the total 160 = 16

=88 participants per group, with a total of 176 participants

### Data collection procedure

Before the study, the researcher organized a training session for 15 research assistants. These include the five residents in obstetrics and gynecology (O/G) and ten midwives. The training session focused on the case selections based on the inclusion and exclusion criteria, measurement of blood pressure using the automated devices and the calibrated sphygmomanometer, administration of oral antihypertensives, monitoring for adverse effects and recording of findings using purpose-designed proforma.

At presentation, Magnesium sulfate was administered to pregnant women according to the Pritchard regimen, which is the hospital protocol. Blood pressure was rechecked after 30 minutes. Those whose blood pressures were still within the severe range (i.e., SBP ≥ 160mmHg and/or DBP ≥ 110mmHg) were recruited for the study after obtaining written informed consent Oxygen saturation (Sp0_2_) was also measured and recorded as the baseline.

The research assistants then opened the sealed, opaque envelope containing the participant’s assigned randomization code number, papers with numbers-1-176, and letter A or B indicating the drug-assigned. The investigator-generated these randomization code numbers. They stated a randomization code number: odd numbers for the Nifedipine group (A) and even numbers for the labetalol group (B), for example-33A for the Nifedipine group and 34B for the labetalol group. Randomization continued until the sample size number was attained.

After the admission of the study participants into the labor ward emergency room, intravenous access was inserted, and blood samples were taken for complete blood count, serum electrolyte, urea, and creatinine; liver function test and clotting profile to know the baseline liver, renal, and hematological status of these women before commencement of treatment. If any derangement was identified, they were managed accordingly. A urethral catheter was passed to monitor urine output. Urinalysis was done using urinalysis strip Medi-Test Combi-3® (Macherey-Nagel GMBH & Co., Germany, Reference number 32031), and the findings were noted. The participants were monitored for MgSO_4_ toxicity after administration of the drug. The respiratory rate, deep tendon reflexes, urine output, and respiratory rate were all monitored. Vials of 10 ml of 10% Calcium gluconate were made available to combat this complication if it was noticed.

Nifedipine was given as an initial dose of 20 mg oral Nifedipine (20 mg extended release Nifedipine (Nifedipine retard [Dexcel® Pharmaceutical] if their systolic blood pressure was ≥ 160 mm Hg and/or their diastolic blood pressure, was ≥ 110 mm Hg one hour after administration of MgSO_4_. An additional 20 mg dose was administered after one hour for two additional doses (to a total of 60 mg) if adequate BP control-SBP <140 mmHg but ≥ 110 mmHg and/or DBP <90 mmHg but ≥ 80 mmHg was not achieved. Administration of further dose was discontinued once adequate blood pressure control was achieved. Pregnant women who were randomly assigned to receive Labetalol were given an initial dose of 200 mg oral labetalol [LABET, Generix® Global Investment Ltd] if their systolic blood pressure was ≥ 160 mm Hg and/or their diastolic blood pressure was ≥ 110 mm Hg. Blood pressure was rechecked after 1 hour. If adequate blood pressure control is not achieved, an additional 200 mg dose can be provided each hour for two additional doses (600 mg).^3^ Adequate blood pressure control was assessed 1 hour after each dose in each group; if adequate blood pressure was obtained, it was adjudged that the primary outcome was achieved for that participant. If not, one of the secondary outcomes was subsequently assessed after the third hour, which was to co-administer the other oral anti-hypertensive, Nifedipine retard for the labetalol group and oral Labetalol for the Nifedipine group as noted in a well-designed purpose oriented proforma. Blood pressure was rechecked every 15 minutes for the next hour, and BP recordings were recorded during the one-hour blood pressure check. If adequate BP control was not achieved afterward, intravenous (IV) anti-hypertensive IV hydralazine was then administered. The dose was administered as recommended by the departmental protocol, which follows ISSHP’s (2018) guidelines [26]. Intravenous Ephedrine at an initial dose of 5-10mg IV bolus and to be repeated every 3-5 minutes to a maximum dose of 30-50mg IV per hour and dopamine infusion at the rate of 2-5microgram/kg/IV infusion to be titrated to effect up to a maximum of 20 microgram/kg/min were made available and within reach to avert severe hypotension. This would be done under the supervision of an obstetrics anesthetist, who would be invited if hypotension was noticed. The time of drug administration was kept and monitored by the investigator and was recorded in the individual participant’s proforma. Intravenous fluid-normal saline was commenced slowly after the third dose so that hypotension would be corrected if detected.

The participant was put on continuous electronic fetal monitor if fetal heart rate irregularity was noticed using the fetal Doppler. The anesthetist, perioperative nurses, and theatre staff were informed and were put on standby so that if fetal heart rate abnormalities were noticed, an emergency cesarean delivery was performed for the participant if intrauterine resuscitation failed to correct fetal heart rate abnormalities. The participants were monitored closely until delivery, during which the maternal and fetal outcomes were further assessed.

Blinding of participants, study investigators, and care providers to group allocation was not possible, given the differences in dose escalation protocols for the two study groups and differences in the color and sizes of the drugs.

The Consort 2010 flow diagram [31] was used for the enrolment, allocation, follow-up of participants in the study, and analysis of the results (Fig. 1). This was illustrated on the next page as a flow diagram of the progress through the phases of a parallel randomized trial of two groups (that is, enrolment, intervention allocation, follow-up, and data analysis):

**Figure 1:**
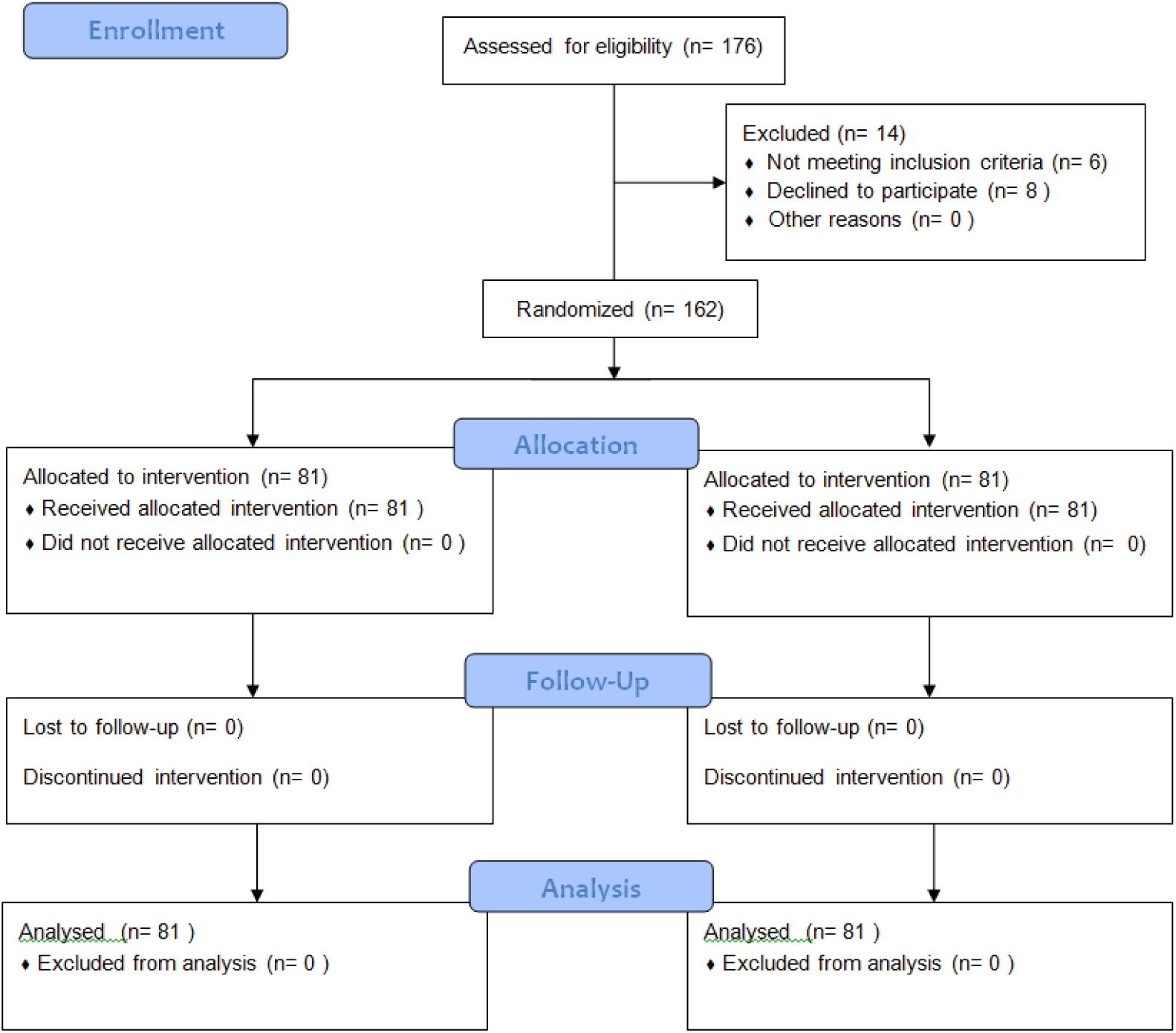
Consort flow diagram for the study

### Outcome Variable Measures

#### Primary Outcome Variable

The primary outcome measure was the percentage of women in both groups who achieved adequate blood pressure control within 6 hours without adverse outcomes.

#### Secondary outcome variables

The secondary outcome measures were the mean BP control in each group, the time between randomization and adequate blood pressure control in each group, the average number of doses used in each group, the Effect of co-administration of both oral antihypertensives on recalcitrant cases, the proportion that required additional antihypertensives in each group, and the adverse outcomes experienced with the administration of oral antihypertensives in each group. The unfavorable outcomes that were sought for included hypotension, visual disturbances, stroke, pulmonary edema (oxygen saturation <90% and abnormal chest x-ray), oliguria (<25 MLS/hours for 2 hours), disseminated intravascular coagulation diagnosed by treating physician, admission to intensive care unit, abruptio placentae, use of fresh frozen plasma, platelet concentrates or fresh whole blood.

### Data Analysis

The sample size was estimated based on prior knowledge of the primary outcome of published data by Easterling et al., which assumed a success rate of 75% (0.75) with Nifedipine T_t_ being the test group and 62.5% (0.625) with labetalol T_C._^3^ being the control group and confidence interval limit at 95% with p-value <0.05, power= 1-B (Type 11 error) at 80%-1-0.8=0.2. Z value at 0.2=0.842. Using the formula for calculating the sample size for a randomized control trial, a sample size of 160 participants was calculated, with 80 participants belonging to each group. The attrition rate of 10% of the total 160 was considered, which put the sample size at 176, 88 participants per group.

Data was collected using a well-designed, purpose-built proforma. The collected data was stored and analyzed using Statistical Package for Social Sciences (SPSS) version 26 for Windows. Intention to treat analysis was done for all the participants.

Categorical variables were analyzed using Chi-square. The percentage success of adequate blood pressure control and the number of doses used in each group were assessed using chi-square. The number and percentage of participants that achieved adequate blood pressure control with co-administration of both antihypertensives and those that needed intravenous hydralazine were evaluated and compared with Chi-square, and statistical significance was determined.

Continuous variables such as blood pressure readings were summarized using mean and standard deviation. The mean and standard deviations of systolic and diastolic blood pressures after each dose of both antihypertensives were compared using a student’s t-test. Fisher’s Exact test was used to assess small frequency counts where the values were less than 5. A p-value of ˂ 0.05 was taken as statistically significant.

#### Ethical Consideration

Ethical clearance was obtained from the hospital’s Research and Ethics Committee. Written informed consent was obtained from willing participants aged 18 years and above who had been counselled on the study while written consent was obtained from the guardians of study participants who are below 18 years old . Participants were informed of their right to withdraw from the study at any point for whatever reason without any consequence on the quality of their antenatal care. Initials and hospital numbers were used to ensure confidentiality. The research was registered under the National Health Research Ethics Committee of Nigeria, Federal Ministry of Health, Nigeria, and Pan African Clinical Trials Registry with trial number **PACTR202406618856160.** Ethics committee approval was obtained from the hospital with ethical approval number **INTERNATIONAL: IRB/IEC/0004553, NATIONAL: NHREC/17/03/2021.**

#### Assessment of Safety

The study did not pose any health risks to the participants.

#### Source of Funding

The investigator funded the study and was involved in its design, data collection, analysis, interpretation, and report writing. The corresponding authors had full access to all the data in the study and were final in deciding whether to submit it for publication.

## Results

### Patient Recruitment

This study screened 176 pregnant women at the two study sites for inclusion. Among the pregnant women eligible for the study, six did not meet the inclusion criteria, and eight declined to participate, making 14 pregnant women excluded. Therefore, 162 women were randomly assigned to the treatment and control groups. Eventually, 81 women were assigned to receive Nifedipine as the treatment group, while 81 women were assigned to receive Labetalol as the control group. None of the women in the two groups were lost to follow-up; neither did they discontinue intervention and were included in the analysis (Fig. 1).

### Demographic Characteristics of the participants

The demographic characteristics of the pregnant women receiving either Nifedipine or Labetalol as part of their treatment are presented (Table 1). There was no statistically significant difference in the age distribution or marital status. Educational status, parity, and booking status between the two groups (p = 0.576, X^2^ = 2.162; p = 0.311, X^2^ = 1.025; p = 0.128, X^2^ = 5.683; p = 0.452, X^2^ = 1.717; p = 0.104, X^2^ = 2.645 respectively).

**Table 1:**
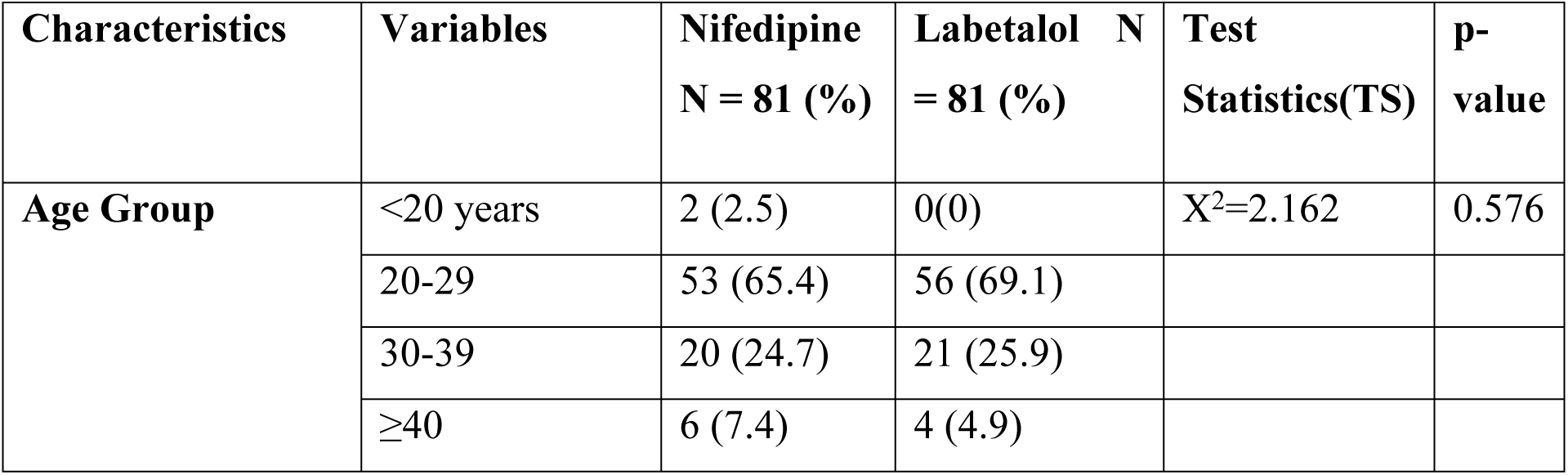
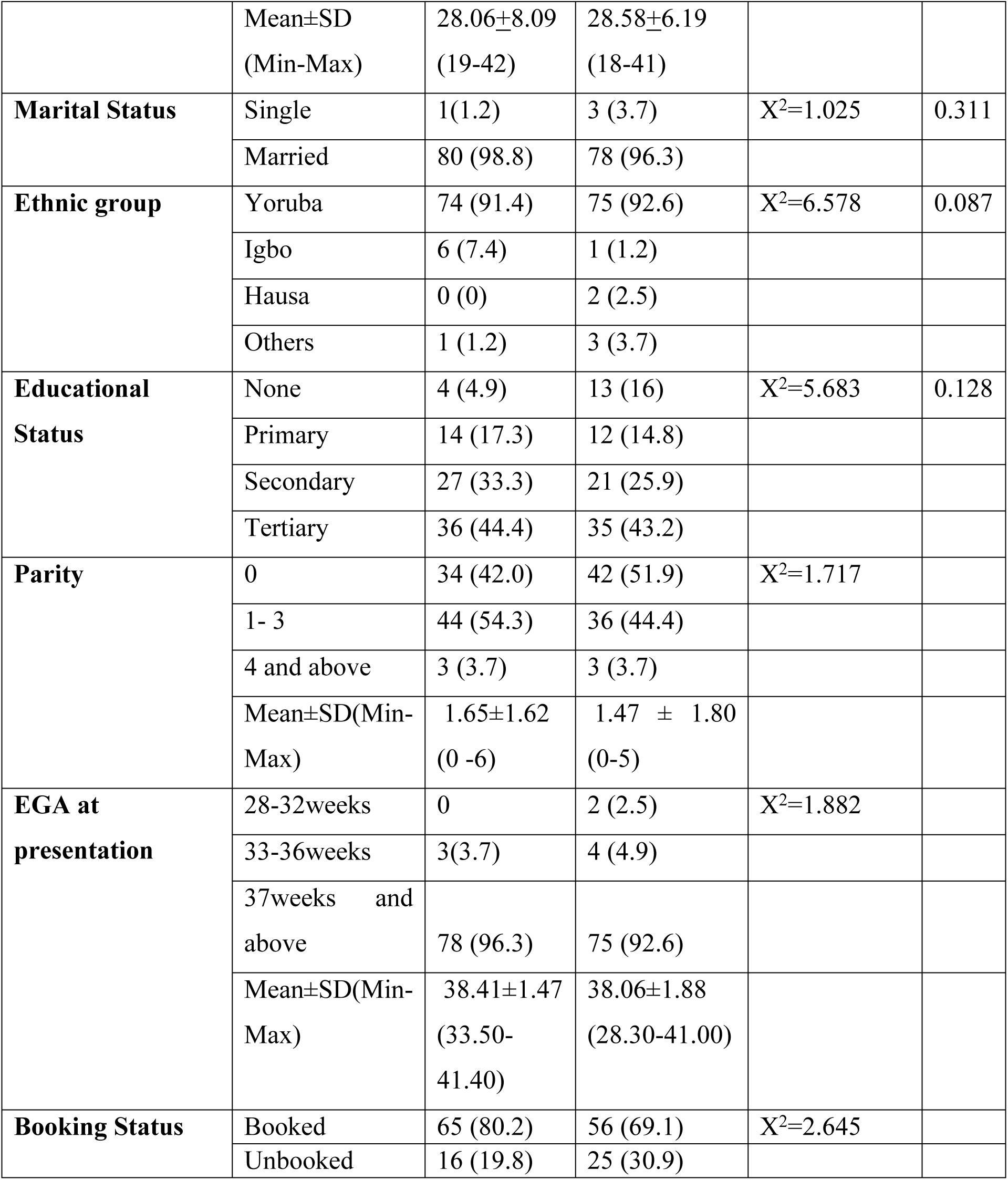
Demographic Characteristics of study participants.

### Demographic Characteristics of Study Participants

#### Baseline Characteristics of the Participants

The baseline characteristics of the participants before the commencement of the interventions are presented in Table 2. There is no statistically significant difference in the mean systolic blood pressure, diastolic blood pressure, oxygen saturation, and heart rate of both groups (p = 0.207, t = -1.266; p = 0.068, t = 1.837; p = 0.405, X^2^ = 0.692; and p = 0.261, X^2^ = –1.129 respectively). However, more women (72% for the Nifedipine group and 76% for the labetalol group) labetalol group had significant proteinuria compared to the Nifedipine group. This difference is statistically significant (p = 0.033, X^2^ = 8.705)

**Table 2:** Baseline Characteristics of Participants.

| Characteristics | Variable | Nifedipine<br>N=81 (%) | Labetalol<br>N= 81(%) | Absolute<br>difference<br>(95% CI) | Test<br>statistics | p-value |
| --- | --- | --- | --- | --- | --- | --- |
| **Systolic Blood Pressure (mmHg)** | <160 | 10 (12.3) | 14 (17.3) |  |  |  |
|  | 160 -179 | 44 (54.3) | 27 (33.3) |  |  |  |
|  | ≥180 | 27 (33.3) | 40 (49.4) |  |  |  |
| | Mean±SD<br>(Min-Max) | 173.51±22.66<br>(150-240) | 178.25±24.95<br>(150-240) | -4.74<br>(-12.14-2.65) | $t = -1.266$ | 0.207 |
| **Diastolic Blood Pressure (mmHg)** | <110 | 20 (23.8) | 24 (29.6) |  |  |  |
|  | 110–119<br>mmHg | 44 (55) | 43 (53.1) |  |  |  |
|  | ≥120<br>mmHg | 17 (21.3) | 14 (17.3) |  |  |  |
| | Mean±SD<br>(Min-Max) | 113±11<br>(90-140) | 110±10<br>(90-140) | 3.099<br>(-0.233–<br>6.431) | $t = 1.837$ | 0.068 |
| | <95 | 2 (2.5) | 4 (4.9) | | $X^2 = 0.692$ | 0.405 |
| **Oxygen saturation<95%** | ≥95 | 79 (97.5) | 77 (95.1) |  |  |  |
| **Heart rate (bpm)** | Mean±SD<br>(Min-Max) | 82.617±10.14<br>(61-104) | 84.407±10.04<br>(62-105) | -1.790<br>(-4.92 – 1.342) | t=-1.129 | 0.261 |
| **ReceivedMgSO4** | Yes | 81 (100) | 81 (100) |  | - | - |
| **Protein in urine** | Negative | 5 (6.2) | 4 (4.9) |  | X<sup>2</sup>=8.705 | 0.033* |
|  | Trace | 6 (7.4) | 7 (8.6) |  |  |  |
|  | 1+ | 11 (13.6) | 8 (9.9) |  |  |  |
|  | 2+ | 12 (14.8) | 18 (22.2) |  |  |  |
|  | ≥3+ | 27 (58.2) | 44 (54.3) |  |  |  |

\* Significant at p<0.05 level

#### Blood Pressure Changes in Response to Treatment

After the first dose of the allocated medication, the mean systolic values of the two groups were 168±27 and 169±30 mmHg for the Nifedipine and Labetalol groups, respectively, with no statistically significant difference (p=0.843, t=-0.198). However, the mean diastolic value of the women in Nifedipine (110±13 mmHg) was significantly higher than that of the Labetalol group (104±10 mmHg) (p=0.004, t=-2.932). More women, 69 (85.2%) in the Nifedipine group received a second of their medication compared to 56 (69.1%) in the labetalol group. This difference was statistically significant (P = 0.015 t= 5.920).

More than half of the women in the Nifedipine (64.2%) group, compared to 23.5% in the Labetalol group, received the third dose of their allocated medication. This difference was statistically significant (p=0.0001, X^2^= 21.627). After the third dose of the allocated medication, the mean diastolic blood pressure values in the Labetalol group (99±5 mmHg) were lower than those in the Nifedipine group. This difference was significant (109±12 mmHg) (p=0.0001, t= -4.391).

The median time between enrolment and adequate blood pressure control for the two groups was 220 minutes in the Nifedipine group and 187 minutes in the labetalol group, with no statistically significant difference between the two groups (p=0.826, Mann Whitney U=3215.000) (Table 3).

**Table 3:** Blood pressure changes in response to treatment.

| Variables |  | Nifedipine<br>n=81 (%) | Labetalol n=81<br>(%) | Absolute<br>difference<br>(95% CI) | Test<br>statistics | p-value |
| --- | --- | --- | --- | --- | --- | --- |
| Median time from recruitment to start of treatment (mins) |  | 31 (8 - 56) | 30 (10 - 50) | 1 (0 -1) | Mann<br>Whitney U-3240.500 | 0.893 |
| Received one dose of allocated medication |  | 81 (100) | 81 (100) |  | - | - |
| Mean Systolic after the first dose (mmHg) | | $168 \pm 27$<br>(120-240) | $169 \pm 30$<br>(120-240) | 0.889<br>(-7.96-9.74) | $t=0.198$ | 0.843 |
| Mean Diastolic after the first dose (mmHg) | | $110 \pm 13$<br>(90-140) | $104 \pm 10$ (90-130) | -5.27<br>(-8.81 - -<br>1.72) | $t=-2.932$ | **0.004\*** |
| Received second dose of allocated medication | | 69 (85.2) | 56 (69.1) | | $X^2=5.920$ | **0.015\*** |
| Mean Systolic after the second dose (mmHg) | | $167 \pm 31$<br>(120-240) | $158 \pm 34$ (120-220) | -8.936<br>(-20.65-2.78) | $t=-1.511$ | 0.134 |
| Mean Diastolic after the second dose (mmHg) |  | 111±12 (90-135) | 103±9 (90-130) | -8.232 (-11.99 - -4.48) | t= -3.419 | **0.001\*** |
| Received the third dose of allocated medication |  | 52 (64.2) | 19 (23.5) |  | X<sup>2</sup>=21.627 | **0.0001\*** |
| Mean Systolic after the third dose (mmHg) |  | 166±37 (120-240) | 150.71±17.51 (122-170) | 10.665 (-7.91 – 29.24) | t= 1.145 | 0.256 |
| Mean Diastolic after the third dose (mmHg) |  | 109±12 (90-135) | 99±5 (90-110) | -9.132 (-12.28 – -4.98) | t= -4.391 | **0.0001\*** |
| Median time between randomization and optimal BP control (mins) |  | 220 (122- 277) | 187 (120-298) | 3.31 (-12.9-19.52) | Mann Whitney U = 3215.000 | 0.826 |
| Received co-administration of another oral anti-hypertensive drug | Labetalol | 23 (28.4) | 0 |  | X<sup>2</sup>=8.984 | **0.003\*** |
|  | Nifedipine | 0 | 16 (19.8) |  |  |  |
| Mean Systolic BP after the co-administration of another oral anti-hypertensive drug |  | 189±41 (124-240) | 163±24 (120-198) | -26.60 (-47.86- -5.35) | t= -2.538 | **0.016\*** |
| Mean Diastolic BP after the co-administration of another oral anti-hypertensive drug |  | 111±11 (90-135) | 100±4 (90-105) | -11.012 (-16.23 – 5.79) | t= -4.318 | **0.0001\*** |
| Received another anti-hypertensive drug during the study period (Hydralazine) |  | 16 (19.6) | 11(13.6) |  | X<sup>2</sup>=0.003 | 0.957 |

\* **Significant at p<0.05 level**

Of the 162 participants, 28.4% (23) in the Nifedipine group were co-administered with Labetalol compared to 19.8% (16) of the women in the Labetalol group who received Nifedipine and this finding was statistically significant (p=0.003, X^2^=8.984) (Table 3). The mean systolic BP after the co-administration of allocated medication for the women in the Nifedipine (A) (189±41mmHg) compared to Labetalol (B) (163±24 mmHg) groups was significantly higher (p=0.016, t= -2.538) (Table 3). In like manner, the mean diastolic blood pressure after the co-administration of allocated medication for the women in the Labetalol (100±4 mmHg) compared to the Nifedipine (111±11mmHg) group was lower with statistical significance. (p=0.0001, t= -4.318) (Table 3).

### Blood pressure changes in response to treatment

#### Percentage Success of blood pressure control, Doses used, Effect of co-administration, and use of intravenous hydralazine

The majority of the participants in the two groups achieved primary outcomes after the third dose without co-administration or additional anti-hypertensive medication (71.6% in the Nifedipine group vs. 80.2% in the labetalol group) with no statistically significant difference between them. (p=0.198, X^2^=1.655) (Table 4). The women in the Labetalol group (30.9%) achieved primary outcome with statistical significance compared to those in the Nifedipine group (14.8%) (p= 0.015, X^2^=5.920) after the first dose of their allocated medication (Table 4). Similarly, after taking the second dose of their allocated medication, more women in the Labetalol group (45.8%) achieved primary outcome with statistical significance compared to those in the Nifedipine group (21%) (p= 0.0001, X^2^=21.627) (Table 4). More women in the Nifedipine group (35.8%) achieved the primary outcome after taking the third dose of their allocated medication with statistical significance (p=0.003, X^2^ = 8.984) (Table 4). After co-administration of the anti-hypertensive therapy, it was observed that 8.6% and 6.2% in the Nifedipine and Labetalol groups, respectively, achieved good blood pressure control with statistical significance between them (p= 0.003, X^2^ = 0.957) (Table 4). The women in the Labetalol group generally achieved primary outcome after 3 hours more than those in the Nifedipine group (80.2% vs. 71.6%) with no significant difference (p=0.198, X^2^ = 1.655) (Table 4). The majority of the participants in both groups reached the blood pressure target (80.2% in the Nifedipine group vs. 86.47% in the Labetalol group) with co-administration of another oral anti-hypertensive agent with no significant difference between the two groups (p=0.292, X^2^ = 1.111) (Table 4).

**Table 4:** Percentage success of blood pressure control, Doses used, Effect of co-administration, and use of intravenous hydralazine.

| Variables | Nifedipine<br>n=81(%) | Labetalol<br>n=81(%) | Test<br>statistics<br>X<sup>2</sup> | p-value |
| --- | --- | --- | --- | --- |
| Achieved primary outcome after three doses. | 58 (71.6) | 65 (80.2) | 1.655 | 0.198 |
| Reached the Blood pressure target | 65 (80.2) | 70 (86.4) | 1.111 | 0.292 |
| Did not reach the blood pressure target | 16 (19.8) | 11 (13.6) |  |  |
| Achieved primary outcome after the first dose | 12 (14.8) | 25 (30.9) | 5.920 | **0.015\*** |
| Achieved primary outcome after the second dose | 17 (21) | 37 (45.8) | 21.627 | **0.0001\*** |
| Achieved primary outcome after the third dose | 29 (35.8) | 3 (3.7) | 8.984 | **0.003\*** |
| Achieved BP target after co-administration of anti-hypeertensive therapy | 7 (8.6) | 5 (6.2) | 0.957 | **0.003\*** |
| Achieved BP target after additional anti-hypeertensive therapy (Hydralazine) | 16 (19.8) | 11 (13.6) |  |  |

**\* Significant at p < 0.05 level**

#### Percentage success of blood pressure control, Doses used, Effect of co-administration, and use of intravenous hydralazine

##### Delivery Outcome

The delivery outcomes of participants in the two groups are presented in Table 5. On the mode of delivery, it was observed that the majority of the women in the two groups delivered via emergency cesarean section (EMCS), with 91.4% and 92.6% for the Labetalol and Nifedipine respectively, with no statistical significance between the mode of delivery of both groups (p=0.772, X^2^= 0.084) (Table 5).

**Table 5:** Delivery Outcome.

| Variables | | Nifedipine<br>n = 81 (%) | Labetalol<br>n = 81 (%) | Test<br>stat -<br>$X^2$ | p-value |
| --- | --- | --- | --- | --- | --- |
| Mode of delivery | SVD | 6 (7.4) | 7 (8.6) | 0.084 | 0.772 |
|  | EMCS | 75 (92.6) | 74 (91.4) |  |  |
| Indication for CS | Severe Chronic Hypertension with superimposed pre-eclampsia and an unfavorable cervix | 18 (24) | 19 (25.7) | 0.056 | 0.813 |
|  | Severe PIH admitted in labor with poor control of blood pressure. | 20 (26.7) | 24 (32.4) | 0.595 | 0.440 |
|  | Pre-eclampsia with severe features and an unfavorable cervix | 23 (30.7) | 20 (27) | 0.24 | 0.624 |
|  | Severe PIH at term (Poorly controlled Blood Pressure) with an unfavorable cervix | 14 (18.6) | 11 (14.9) | 0.386 | 0.535 |

\* Significant at $p<0.05$ level

Fourteen of the participants in the Nifedipine group vs. 11 (14.9%) in the Labetalol group had EMCS due to severe PIH (poorly controlled Blood Pressure), with no significant difference between the two groups (p=0.535, X^2^=0.386) (Table 5).

##### Delivery Outcome

SVD: Spontaneous Vaginal Delivery; EMCS: Emergency Caesarean Section; PIH: Pregnancy Induced Hypertension

##### Fetal Outcome

Most of the women were admitted at a mean gestational age of 38.41± 1.47 weeks for the Nifedipine group and 38.06±1.88 weeks for the labetalol group. Most of the babies were delivered at term with the average estimated gestational age of 39.06±1.38 for the Nifedipine group and 39.07±1.40 weeks for the labetalol groups, respectively, with no significant difference (p=0.955, t = -0.056) (Table 6). The outcome of delivery showed no stillbirth in the Nifedipine group compared with one (4.9%) preterm stillbirth in the Labetalol group with no statistically significant difference (p = 1.006, X^2^=4.101). A perinatal death was observed only in the Labetalol group before discharge (Table 6).

**Table 6:** Fetal Outcome.

| Variables |  | Nifedipine<br>n = 81 (%) | Labetalol<br>n = 81 (%) | Absolute<br>difference<br>(95% CI) | p-value | Test<br>statistics |
| --- | --- | --- | --- | --- | --- | --- |
| EGA at<br>delivery(weeks) | | $39.06 \pm 1.38$ | $39.07 \pm 1.40$ | $-0.01(-0.44-0.42)$ | 0.955 | $t= -0.056$ |
| Outcome of<br>Delivery | Live birth | 81 (100) | 80 (98.8) | | 1.006 | $X^2=4.101$ |
|  | Stillbirth | 0 | 1 (1.2) |  |  |  |
| Birth weight | Mean±SD (kg) | $2.97 \pm 0.75$ | $3 \pm 0.38$ | $0.04 (-0.15 - 0.22)$ | 0.694 | $t= 0.394$ |
|  | Preterm ELBW | 0 | 0 |  |  |  |
|  | Preterm VLBW | 0 | 2(2.5) |  |  |  |
|  | Low Birth<br>Weight | 8(9.9) | 10(12.3) |  |  |  |
|  | Normal Birth<br>Weight | 73 (90.1) | 69(85.2) |  |  |  |
| APGAR Score at 5 minutes (+) | <7 | 11 (13.6) | 8 (9.9) |  | 0.481 | X<sup>2</sup>=0.496 |
|  | 7 and above | 70 (86.4) | 72 (88.9) |  |  |  |
| Incubated at the place of delivery |  | 3(3.7) | 1 (1.2) |  | 0.316 | X<sup>2</sup>=1.006 |
| Need for NICU admission |  | 11 (13.6) | 9 (11.1) |  | 0.386 | X<sup>2</sup>=0.753 |
| Reason for NICU admission | Birth Asphyxia | 9(81.8) | 8 (88.9) |  | 0.481 | X<sup>2</sup>=0.496 |
|  | Prematurity | 4(36.4) | 5(62.5) |  | 0.746 | X<sup>2</sup>=0.131 |
|  | Tachypnoea of the newborn | 3(27.3) | 2(25) |  | 0.506 | X<sup>2</sup>=0.194 |
| Birth Asphyxia | Mild | 6 (66.7) | 4 (50) |  | 0.660 | X<sup>2</sup>=1.382 |
|  | Moderate | 2 (22.2) | 3 (37.5) |  |  |  |
|  | Severe | 1 (11.1) | 1(12.5) |  |  |  |
| Perinatal death before mother's discharge | Yes<br>No | 0(0)<br>81 (100.0) | 1(1.25)<br>79 (98.75) |  | 0.316 | X<sup>2</sup>=1.006 |

\* Significant at $p < 0.05$ level
NICU: Neonatal Intensive Care Unit

##### Fetal Outcome

###### Maternal Outcome

The result of the maternal outcome is presented in Table 7. The result showed that 2 (2.5%) and 4 (4.9%) of the participants in the Labetalol and Nifedipine groups, respectively, had adverse maternal outcomes with no significant difference between the two groups (p = 0.405, Fischer Exact = 0.692) (Table 7). One (1, 25%) of the participants in the labetalol group vs. none in the Nifedipine group had abruptio placentae complicated by disseminated intravascular coagulopathy and was admitted to ICU. This participant later died at the ICU on account of complications of the disease not from intervention instituted during the study. This was the only maternal mortality recorded in all the participants. There was no statistically significant difference (p = 0.316, Fischer Exact = 1.006) (Table 7).

**Table 7:**
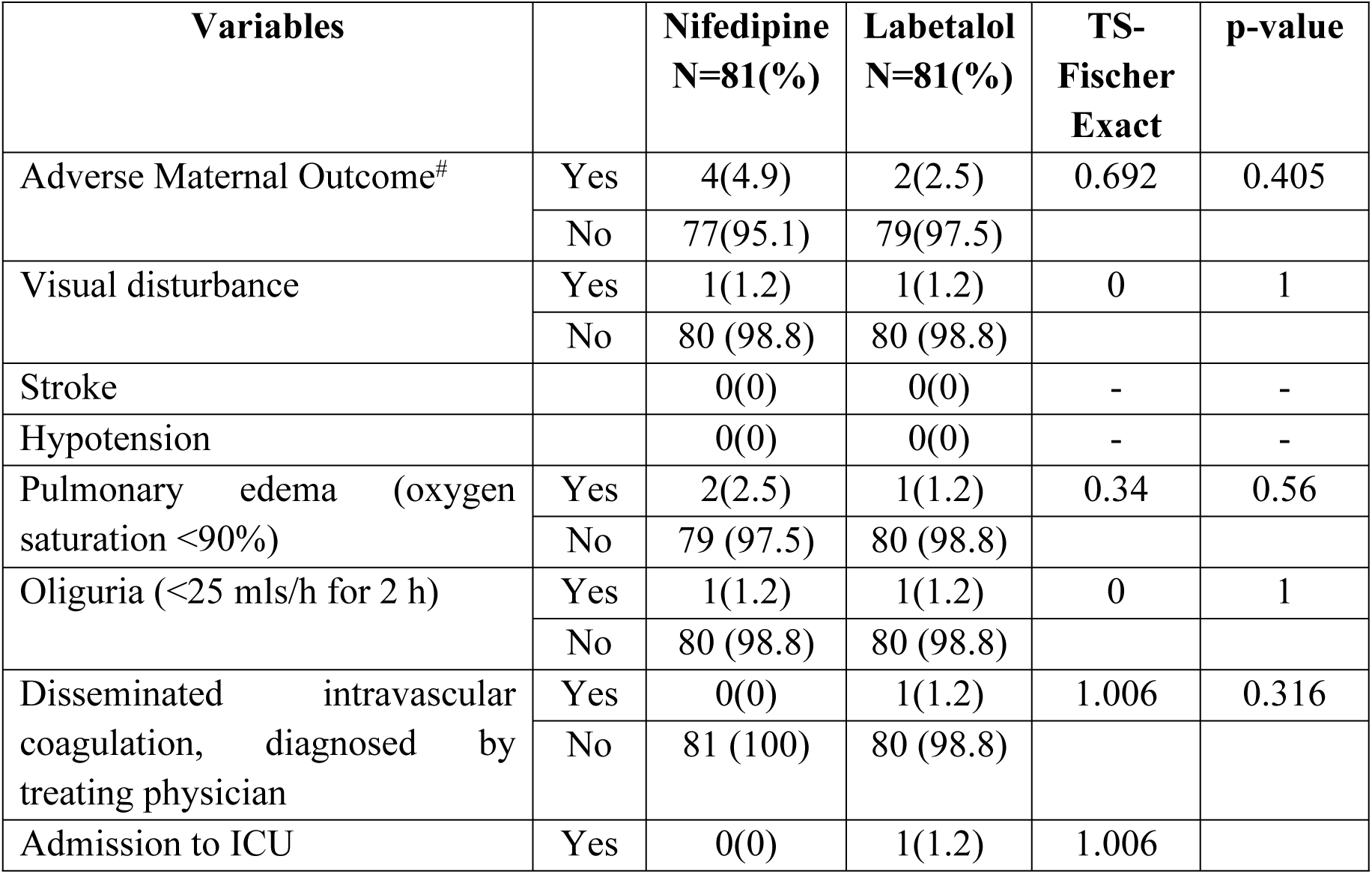
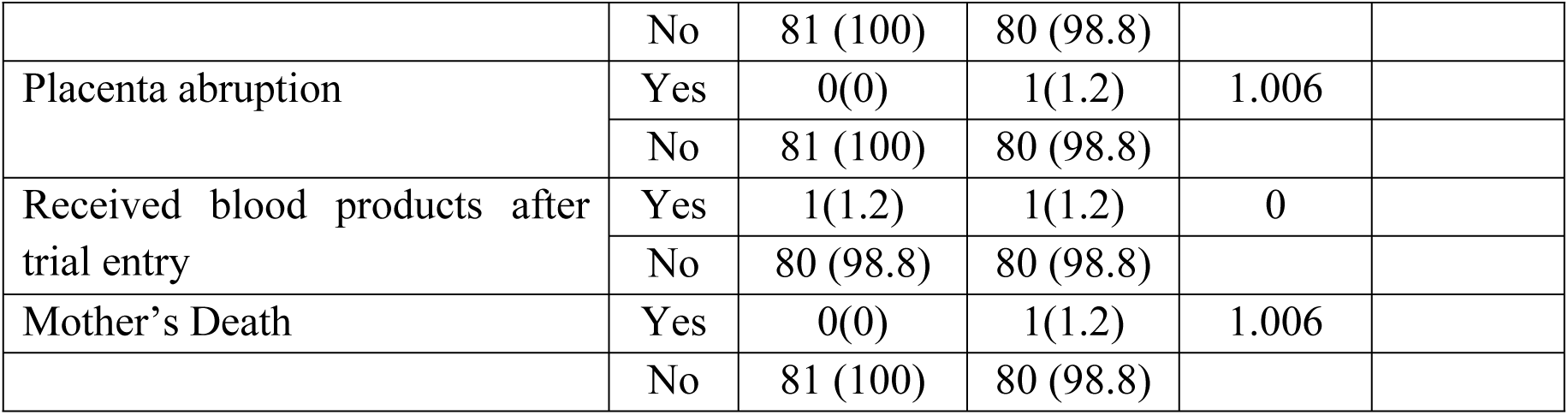
MATERNAL OUTCOME.

## Discussion

Despite recent advances and studies in hypertensive disorders in pregnancy, it is still a significant cause of maternal and perinatal mortality and morbidity [32]. Management should depend on the severity of Hypertension. Besides delivery of the fetus, the primary treatment of severe hypertensive disorders of pregnancy at term or close to term is control of blood pressure with the use of anti-hypertensive drugs [1, 32]. Specific anti-hypertensive agents are used to prevent and treat severe Hypertension, most of which are given by the parenteral route as recommended by most guidelines [1, 4, 32]. These require an intravenous route to administer, which may not be readily available in low-resource settings. A study on the efficacy of oral anti-hypertensive drugs in controlling high blood pressure is essential to determine the first-line oral anti-hypertensive drug to control blood pressure for stocking and institute prompt treatment to prevent complications both for women and fetuses [33].

Our study revealed that more participants in the labetalol group achieved adequate blood pressure control compared to the Nifedipine group though, there is no statistical difference between them. This was comparable to the findings in the studies by Deshmukh et al. [33] and van de Vusse et al. [34] but in contrast with the findings of Easterling et al. [2], which showed that the Nifedipine group had better control of blood pressure than the Labetalol and the alpha methyldopa group. The index study had adequate blood pressure control at the normal range of blood pressure control, which differed from the cut-off values used by the Adebayo et al. study [35], which targeted adequate control at SBP <150 mmHg and DBP < 100 mmHg, which was still in the Hypertension range. Findings in the index study and study by Adebayo et al. were comparable concerning maternal and fetal outcomes, which showed that adequate blood pressure control within normal ranges should be aimed at during management to further reduce the risk of maternal complications as a result of poor blood pressure control.

There was a significant reduction in the mean systolic BP and diastolic BP after administering the first, second, and third doses of oral medication in the Labetalol (B) group compared to the Nifedipine (A) group. This was contrary to the findings in the study by Easterling et al., where the reverse was the case [2]. The proportion of participants that received the second and third doses of the antihypertensives was lesser in the labetalol group. This was similar to that seen in a study by Deshmukh et al. [33]. In a systemic review conducted on oral anti-hypertensive therapy for the management of severe Hypertension in pregnancy by George et al [36], it was found that oral Nifedipine, Labetalol, and methyldopa are suitable options for the treatment of severe Hypertension in pregnancy/postpartum. In another meta-analysis conducted to compare the efficacy and safety of Nifedipine with other anti-hypertensive medications for the management of hypertensive disorders in pregnancy by Wu et al., Nifedipine was found to be more effective than other anti-hypertensive medications in reducing blood pressure, particularly in patients with severe Hypertension [16].

The participants in the Labetalol group achieved adequate BP control earlier than those in the Nifedipine group even though the difference was not statistically significant. This finding is similar to that of Deshmukh et al. [33]

More participants in Nifedipine to Labetalol group had co-administration of another anti-hypertensive labetalol in a bid to control Hypertension despite previous use of 3 doses of Nifedipine. This observation is comparable to the finding in the Deshmukh et al. study [33]. More participants in the Nifedipine group required the use of intravenous hydralazine when compared to group B, which was similar to findings by Deshmukh et al. [33]

Overall, the Labetalol group had better BP control with fewer drug doses. This was contrary to the findings of Easterling et al. [2]. The exact reason for this discrepancy is not known. However this may be due to difference in geographical location. Co-administration of both antihypertensives improved the control of severe Hypertension among participants with recalcitrant blood pressure control. This is compared favorably with the findings of previous studies, which showed that this is a practice worth adopting [37–39].

Maternal adverse outcomes and side effect profiles were more common in the Nifedipine group compared to the Labetalol group, though this was not statistically significant. This compared favorably with previous studies [40–41]

### Study limitations

The limitations of our study are that the study population included pregnant women with both proteinuric and non-proteinuric chronic and gestational severe Hypertension. This may have affected blood pressure control, as it was noticed that it was easier to control Hypertension in pregnant women with non-proteinuric severe Hypertension than those with proteinuric severe Hypertension.

Blinding was not employed in the study. This is because the different doses, colors, and sizes of both drugs made it difficult to mask them. This drawback may have increased the risk of bias.

## IMPLICATION OF THE STUDY

The study confirmed the efficacy of two common oral antihypertensives in treating severe Hypertension in pregnancy. These medications are readily available and do not require any expertise to administer. Adverse effects and side effects on the mother or the fetus are less compared to the intravenous regimen.

Furthermore, because of the first-pass Effect, the severity of adverse effects is less than that of intravenous medications. By prompt institution of treatment with administration of readily accessible oral antihypertensives, the maternal and fetal complications posed by uncontrolled Hypertension can be reduced or avoided.

Determining the relative efficacy between the two on a larger scale will encourage stocking the more effective drug and achieving adequate blood pressure control with fewer doses, less time, and minimal adverse fetal outcomes.

## Conclusion

Both oral antihypertensives were effective in the control of severe Hypertension in pregnancy in our study location. This study showed that oral Labetalol had better control of severe Hypertension with fewer doses of the drug and a shorter duration of time, especially when the systolic BP and/or DBP were < 180 mmHg and < 120 mmHg, respectively, compared to oral Nifedipine. Nifedipine from the study showed that SBP and DBP above this range were not controlled with both medications. Therefore, it may be preferable to use intravenous hydralazine to control BP above the ranges above. However, further studies are needed to corroborate this assertion. Both oral antihypertensives should be encouraged with a preference towards using oral Labetalol as the first line in rural or sub-urban health centers to control severe Hypertension before transfer to secondary and tertiary centers to reduce perinatal and maternal morbidity or mortality. Larger scale studies are required to establish the role of oral anti-hypertensive in the management of severe Hypertension in pregnancy in our locality.

## Data Availability

All relevant data are within the manuscript and its Supporting Information files.

## Acknowledgment

We sincerely thank the staff of the Obafemi Awolowo University Teaching Hospitals Complex, the Obafemi Awolowo University, and all the study participants for their support.

## Authors contribution

**Conceptualisation**: Obinna C Eze

**Data curation:** Obinna C Eze, Akintunde O Fehintola, Akinfaderin D. Adeyinka

**Formal analysis:** Obinna C Eze, Clement A Adepiti, Ibraheem O Awowole, Adebimpe O Ijarotimi, Babalola E. Olajide

**Investigation:** Obinna C Eze, Akintunde O Fehintola, Ajiboye D Akinyosoye, Abdur-Rahim F Zainab, Ayegbusi E Oluwole, Ernest O Orji

**Methodology:** Obinna C Eze, Akintunde O Fehintola, Akinfaderin D. Adeyinka, Ernest O Orji

**Project Administration:** Obinna C Eze

**Supervision:** Akintunde O Fehintola, Akinfaderin D. Adeyinka, Ernest O Orji

**Validation**: : Obinna C Eze, Clement A Adepiti, Ibraheem O Awowole, Adebimpe O Ijarotimi, Babalola E. Olajide

**Writing-original draft:** Obinna C Eze

**Writing-review & editing:** Obinna C Eze, Akintunde O Fehintola, Ajiboye D Akinyosoye, Abdur-Rahim F Zainab, Ayegbusi E Oluwole, Ernest O Orji

## References

1. Tranquilli A, Dekker G, Magee L, Roberts J, Sibai B, Steyn W. The classification, diagnosis and management of the hypertensive disorders of pregnancy: a revised statement from the ISSHP. Pregnancy hypertension 2014; 4: 97.

2. Easterling T, Mundle S, Bracken H, Parvekar S, Mool S, Magee LA. Oral anti-hypeertensive regimens (nifedipine retard, Labetalol, and methyldopa) for management of severe Hypertension in pregnancy: an open-label, randomized controlled trial. Lancet 2019; 394: 1011–21

3. Hypertension in pregnancy: diagnosis and management: nice guidance 2018.nice.uk.org/guidance/CG107; 3-10

4. Webster LM, Myers JE, Nelson-Piercy C, Harding K, Cruickshank JK, Coote I. Labetalol Versus Nifedipine as Antihypertensive Treatment for Chronic Hypertension in Pregnancy; A Randomized Controlled Trial: Hypertension 2017 Nov;70(5):915-922. https://www.ahajournals.org/doi/10.1161/HYPERTENSIONAHA.117.09972

5. Uzan J, Carbonnel M, Piconne O, Asmar R and Ayoubi J-M. Pre-eclampsia: pathophysiology, diagnosis, and management. Vasc Health Risk Manag 2011; 7: 467.

6. Koopmans CM, Bijlenga D, Groen H, Vijgen SM, Aarnoudse JG, Bekedam DJ, et al. Induction of labour versus expectant monitoring for gestational Hypertension or mild pre-eclampsia after 36 weeks’ gestation (HYPITAT): a multicentre, open-label randomized controlled trial. The Lancet 2009; 374: 979–988.

7. Podymow T, August P. Update on the use of antihypertensives in pregnancy. DOI: 10.1161/HYPERTENSIONAHA.106.075895 2008;51: 960–969

8. Shennan A, Waugh J. The measurement of blood pressure and proteinuria in pregnancy. Pre-eclampsia. RCOG Press, London, England, 2003, pp.305–324.

9. Pickles CJ, Symonds EM, Pipkin FB. The fetal outcome in a randomized, double-blind controlled trial of Labetalol versus placebo in pregnancy-induced Hypertension. Br J Obstet Gynaecol. 1989; 96: 38–43. Cross ref Medline Google Scholar

10. Magee LA, Schick B, Donnenfeld AE, Sage SR, Conover B, Cook L, McElhatton PR, Schmidt MA, Koren G. The safety of calcium channel blockers in human pregnancy: a prospective, multicenter cohort study. Am J Obstet Gynecol. 1996; 174: 823– 828.CrossrefMedlineGoogle Scholar

11. Bartels PA, Hanff LM, Mathot RA, Steegers EA, Vulto AG, Visser W. Nicardipine in pre-eclamptic patients: placental transfer and disposition in breast milk. BJOG. 2007;114:230 –233.

12. Jannet D, Carbonne B, Sebban E, Milliez J. Nicardipine versus metoprolol in the treatment of Hypertension during pregnancy: a randomized comparative trial. Obstet Gynecol

13. Wide-Swensson DH, Ingemarsson I, Lunell NO, Forman A, Skajaa K, Lindberg B, Lindeberg S, Marsal K, Andersson KE. Calcium channel blockade (isradipine) in treatment of Hypertension in pregnancy: a randomized placebo-controlled study. Am J Obstet Gynecol. 1995;173: 872– 878

14. Ales K. Magnesium plus nifedipine. Am J Obstet Gynecol. 1990; 162: 288.Google Scholar

15. Puzey MS, Ackovic KL, Lindow SW, Gonin R. The Effect of nifedipine Nifedipinembilical artery Doppler waveforms in pregnancies complicated by Hypertension. S Afr Med J. 1991; 79: 192–194.MedlineGoogle Scholar

16. Brown MA, Buddle ML, Farrell T, Davis GK. Efficacy and safety of nifedipine tablets for the acute treatment of severe Hypertension in pregnancy. Am J Obstet Gynecol. 2002; 187: 1046–1050.CrossrefMedlineGoogle Scholar

17. Ben-Ami M, Giladi Y, Shalev E. The combination of magnesium sulfate and nifedipine: Nifedipine for neuromuscular blockade. Br J Obstet Gynaecol. 1994; 101: 262– 263.CrossrefMedlineGoogle Scholar

18. Waisman GD, Mayorga LM, Camera MI, Vignolo CA, Martinotti A. Magnesium plus nifedipine: Nifedipineion of hypotensive Effect in preeclampsia? Am J Obstet Gynecol. 1988; 159: 308–309.CrossrefMedlineGoogle Scholar

19. Altman D, Carroli G, Farrell B, Moodley J, Neilson J, Smith D. Do women with pre-eclampsia, and their babies, benefit from magnesium sulphate? The Magpie Trial: a randomized placebo-controlled trial. Lancet. 2002; 359: 1877– 1890.CrossrefMedlineGoogle Scholar

20. Duley L, Henderson-Smart DJ, Meher S. Drugs for treatment of very high blood pressure during pregnancy. Cochrane Database Syst Rev. 2006;3:CD001449.

21. Sibai BM. Diagnosis and management of gestational Hypertension and preeclampsia. Obst & Gyn. 2003; 102(1):181–92.

22. Scardo JA, Vermillion ST, Hogg BB, Newman RB. Hemodynamic effects of oral nifedipine Nifedipinemptic hypertensive emergencies. Am J Obstet Gynecol. 1996; 175: 336 338;discussion 338–340.CrossrefMedlineGoogle Scholar

23. Lindow SW, Davies N, Davey DA, Smith JA. The Effect of Sublingual Nifedipine on Placential Blood Flow in Hypertensive Pregnancy. Br J Obstet Gynaecol. 1988;95:1276–1281.

24. Kenny LC. Hypertensive Disorders of Pregnancy. In: Kenny LC and Meyers JE (eds) Obstetrics by Ten Teachers. 20th ed. Broken Sound Parkway NW: CRC Press, 2017, 133–135

25. Arulkumaran N, Lightstone L. Severe pre-eclampsia and hypertensive crises. Best Pract Res Clin Obstet Gynaecol 2013; 27: 877–884.

26. Sibai BM. Diagnosis and management of gestational Hypertension and preeclampsia. Obst & Gyn. 2003; 102(1):181–92.

27. Makinde ON, Ezechi O (Ed.): The Contribution of Severe Pre-Eclampsia and Eclampsia to Perinatal Mortality in a Nigerian Teaching Hospital, Perinatal Mortality, https://www.intechopen.com/books/765. 2012; 111–117

28. Baoliang Zhong MD: How to calculate sample size in randomized controlled trial; J Thorac Dis. 2009 Dec; 1(1): 51–54

29. Wittes J. Sample size calculations for randomized controlled trials; Epidemiologic reviews by John Hopkins Bloomberg School of Public Health; 2002;24(1): 39–44

30. The consort flow diagram. Transparent reporting of trials. https://www.consort-statement.org/download/Media/Default/Downloads/CONSORT%202010%20Flow%20Diagram.doc

31. Magee LA, Smith GN, Bloch C, et al. Guideline No. 426: Hypertensive Disorders of Pregnancy: Diagnosis, Prediction, Prevention, and Management. J Obstet Gynaecol Can. 2022;44(5):547–571.e1. doi:10.1016/j.jogc.2022.03.002

32. Shekhar S, Sharma C. Oral Nifedipine or intravenous Labetalol for hypertensive emergency in pregnancy: a randomized control trial. Obstet Gynecol. 2013;122(5):1057–63.

33. Deshmukh UB, Savitha A, Tengli S. Comparative study of Labetalol and nifedipine Nifedipineent of hypertensive disorders of pregnancy in BRIMS tertiary care center. The New Indian Journal of OBGYN. 2021; 8(1): 117–20.

34. Stott D, Bolten M, Salman M, Paraschiv D, Douiri A, Kametas NA. A prediction model for the response to oral Labetalol for the treatment of antenatal Hypertension. Journal of Human Hypertension, 2016; 31(2): 126–31.

35. Adebayo J, Nwafor JI, Lawani LO, Esike CO. Efficacy of nifedipine and hydralazine in the management of severe Hypertension in pregnancy: A randomized controlled trial. Nigeria postgrad med jour., Oct-Dec 2020;27(4):317–324. doi: 10.4103/npmj.npmj_275_20.

36. Firoz T, Magee LA, MacDonell K, Payne BA, Gordon R, M Vidler. Community Level Interventions for Pre-eclampsia (CLIP) Working Group: a systematic review on Oral antihypertensive therapy for severe Hypertension in pregnancy and postpartum: DOI: 10.1111/1471-0528.12737 www.bjog.org, 2014.

37. George R, Thomas C, Joy CA, Varghese B, Undela K, Adela R. Comparative efficacy and safety of oral nifedipine Nifedipine antihypertensive medications in the management of hypertensive disorders of pregnancy: a systematic review and meta-analysis of randomized controlled trials. Journal of Hypertension. 2022 Oct 1;40(10):1876–86.

38. Kilpatrick SJ, Abreo A, Greene N, Melsop K, Peterson N, Shields LE, Main EK. Severe maternal morbidity in a large cohort of women with acute severe intrapartum Hypertension. American journal of obstetrics and gynecology. 2016 Jul 1;215(1):91–e1.

39. Hangarga US, Rita D, Harshitha K. Comparative study of Labetalol and Nifedipine in management of hypertensive disorders in pregnancy. Int J Reprod Obstet Gynecol. 2017;6:194–7.

40. Rose DT, Jeyarani P. Comparative study of Labetalol and nifedipine Nifedipineent of non-severe preeclampsia and its fetomaternal outcome. Int J Reprod Contracept Obstet Gynecol. 2019;8(5):2035

41. Patel AR, Arora SR, Bhatt JK. Comparison of oral nifedipine Nifedipineabetalol as a single drug therapy for control of blood pressure in preeclampsia. Int J Reprod Contracept Obstet Gynecol 2020;9:2328–32

